# Knowledge and utilization of family planning and safe abortion services among married women of reproductive age in the Raute community of Nepal: a census-based cross-sectional study

**DOI:** 10.64898/2026.05.18.26353535

**Authors:** Manoj Joshi, Anjali Bhatt, Anusha Sharma, Milan Thapa, Sudip Khanal

## Abstract

Indigenous and nomadic communities worldwide face disproportionate and persistent barriers to reproductive health services, including family planning and safe abortion. The Raute of Nepal — one of the country’s last nomadic hunter-gatherer groups represent a uniquely marginalized population for whom no prior population-level quantitative reproductive health data exist. This gap prevents health authorities and program implementers from designing evidence-based, culturally appropriate interventions for this community. This census-based cross-sectional study enrolled all 192 eligible married women of reproductive age in the Raute community of Parshuram Municipality, Dadeldhura district, Sudurpaschim Province, Nepal. Data were collected through structured, pre-tested, face-to-face interviews, and analyzed using descriptive statistics, chi-square tests, and binary logistic regression in IBM SPSS version 16. More than half of participants (53.6%) currently used family planning, with injectable contraceptives being the most common method (42.7%), followed by female sterilization (33.0%) and implants (24.3%). Condom use was negligible at 1.0%. Among non-users (46.4%), 97.7% cited lack of interest as the primary reason for non-use. Knowledge of safe abortion services was reported by 61.5% of women, yet only 8.3% had ever accessed such services, and awareness of Nepal’s national safe abortion policy, which has been in effect since 2002 was critically low at 10.4%. In bivariate analysis, no socio-demographic or socioeconomic variable was significantly associated with family planning use. The sole significant independent predictor of current family planning utilization in the adjusted logistic regression model was non-utilization of safe abortion services (adjusted odds ratio = 4.275; 95% confidence interval: 1.145–15.954; p = 0.030), suggesting that contraceptive use and abortion service use represent alternative reproductive management strategies in this community. Younger age (≤30 years) and urban residence were significantly associated with safe abortion use in bivariate analysis but were attenuated after adjustment, reflecting limited statistical power arising from the small number of outcome events (n = 16). These findings reveal critical gaps in reproductive method diversity, safe abortion policy literacy, and male partner engagement. Community-based mobile outreach tailored to nomadic movement patterns, targeted legal literacy programs in the local language, and structured male involvement strategies are urgently required to improve reproductive health equity in this vulnerable indigenous population.

## Introduction

Reproductive health is a fundamental human right, encompassing the capacity of individuals and couples to decide freely whether, when, and how many children to have [1]. Family planning and safe abortion are two intersecting pillars of this right: the former enables individuals to control the timing and spacing of pregnancies through modern contraception, while the latter provides a safe option when contraception fails or is unavailable. Together, these services are critical to reducing maternal mortality, preventing unintended pregnancies, and advancing gender equity [2, 3]. Globally, more than 1.1 billion women of reproductive age required family planning services in 2021, and 164 million had an unmet need for contraception [4]. An estimated 45% of all abortions worldwide are unsafe, with the burden concentrated in low- and middle-income countries and among the most marginalized women [5]. The gap between reproductive health policy commitments and service delivery realities is most acute for indigenous, nomadic, and rural communities whose needs are systematically underrepresented in national surveys and underserved by formal health systems.

Indigenous peoples, numbering approximately 476 million worldwide and comprising around 6% of the global population, bear a disproportionate burden of reproductive ill-health [6]. Indigenous women experience maternal mortality ratios two to three times higher than national averages in many countries [7, 8]. High fertility rates, short birth intervals, early marriage, undernutrition, and physically demanding labor during pregnancy amplify obstetric risk, while geographic remoteness, linguistic marginalization, cultural incompatibility of health services, poverty, and distrust of state institutions compound barriers to care [9, 10]. Family planning programs designed for settled populations and fixed facilities fail to reach nomadic communities whose movement patterns are incompatible with scheduled outreach. Safe abortion services present additional challenges given stigma, limited facility access, and constrained reproductive autonomy [5, 11].

In Nepal, the government has set modern contraceptive prevalence targets of 53% by 2022 and 60% by 2030 under Sustainable Development Goal 3.7.1(a). The Nepal Demographic and Health Survey (NDHS) 2022 reported a contraceptive prevalence rate of 57% among currently married women, yet 21% still have an unmet need and 78% express demand for family planning [12]. These national figures mask profound disparities: indigenous and marginalized ethnic groups including Dalits, Muslims, and Janajatis consistently demonstrate the lowest contraceptive prevalence, with ethnicity identified as a significant predictor of unmet need in multiple national studies [13, 14]. Nepal legalized abortion in 2002 under a progressive framework permitting termination on request up to 12 weeks, up to 18 weeks in cases of rape or incest, and at any gestation when the woman’s life or health is at risk [15]. Despite this, awareness of the law and utilization of safe abortion services remain substantially lower among women from marginalized communities [16, 17]. A study among Tharu indigenous women of Dang district reported a 49% unmet need for family planning [18], illustrating the broader reproductive health deficit among Nepal’s indigenous populations.

The Raute community sits at the extreme end of this vulnerability spectrum. Widely regarded as one of Nepal’s last nomadic hunter-gatherer groups, they maintain a way of life that has remained largely unchanged for generations. According to the 2021 National Census, the total Raute population stands at 566 individuals (289 males, 277 females), a figure reflecting a documented decline attributed to premature deaths from malnutrition, inadequate healthcare, poor shelter, and near-complete absence of reproductive health services [19, 20]. The Raute govern themselves through a traditional leadership system called “Mukhiya,” travel seasonally across western Nepal, and subsist through primate hunting, forest foraging, and trade of hand-carved wooden goods [19]. Traditional healers remain the first point of contact for most health complaints, and deep-rooted fatalistic beliefs substantially limit health-seeking behavior [21]. All women in the community are homemakers; 54.2% married at or before age 16, well below Nepal’s legal marriage age of 20 [22]. Prior qualitative research documented near-absent antenatal care attendance and forest deliveries among Raute women in Dailekh district, driven by distance, cultural norms, and transport absence [21]. Despite this evidence base, no peer-reviewed quantitative study has assessed family planning or safe abortion service utilization among Raute women at the population level, leaving a critical evidence gap for health system planning and programmatic design.

The present study was conducted to fill this gap by providing the first population-level data on family planning and safe abortion knowledge and utilization among married Raute women of reproductive age, and by identifying factors independently associated with service use. Specifically, the study aimed to: (1) describe the prevalence and patterns of family planning utilization, including method type, duration, perceived advantages, and reasons for non-use; (2) describe safe abortion service knowledge and utilization, including national policy awareness; (3) examine bivariate associations between socio-demographic and socioeconomic variables and both outcomes; and (4) identify independent predictors of both outcomes using logistic regression analysis. The findings are intended to inform local government health planning, NGO programming, and national indigenous reproductive health policy.

## Materials and methods

### Ethics statement

This study was conducted in accordance with the Declaration of Helsinki. Ethical approval was obtained from the Institutional Review Committee of Manmohan Memorial Institute of Health Sciences (Reference: MMIHS-IRC 886; 1 September 2022). Prior written permission was secured from the health section of Parshuram Municipality, Dadeldhura district. All participants received a full explanation of the study purpose, procedures, voluntary nature of participation, and their right to withdraw at any time without consequence before enrolment. Written informed consent was obtained from each participant prior to interview. No personally identifying information was recorded in the questionnaire, ensuring participant anonymity and data confidentiality throughout the study. This manuscript is reported in accordance with the Strengthening the Reporting of Observational Studies in Epidemiology (STROBE) guidelines for cross-sectional studies.

### Study design, setting, and population

A census-based cross-sectional descriptive and analytical study was conducted in Parshuram Municipality, Dadeldhura district, Sudurpaschim Province, Nepal. The municipality was selected because it represented the current primary residence of the Raute community at the time of the study. At the time of data collection, the total Raute population in the municipality numbered 419 individuals, of whom 192 were married women of reproductive age (15–49 years). A census method was adopted, enrolling all 192 eligible women. Census sampling was employed rather than probability sampling because: (a) the total eligible population was small and fully accessible; (b) it eliminated selection bias; and (c) it maximized statistical power for outcomes with low event rates, such as safe abortion service use. No formal sample size calculation was performed, as census enumeration of the entire eligible population was feasible and appropriate. Women of reproductive age who were unmarried or who had a reported mental disability were excluded. Data collection commenced on 25 September 2022, following ethical approval, and ended on 23 November 2022.

### Data collection tool and procedures

A structured questionnaire was developed through systematic review of published literature and adapted to the local context. No validated instrument specific to this population was available; the tool was therefore purpose-built and reviewed for content validity by the research supervisor and departmental faculty. The instrument comprised three sections: (a) socio-demographic and socioeconomic characteristics; (b) family planning knowledge, method type, duration of use, perceived advantages, source of services, and reasons for non-use; and (c) safe abortion knowledge, service location awareness, utilization history, type of procedure received, and knowledge of Nepal’s national safe abortion policy. To assess reliability, the tool was pre-tested on 10% of women (n ≈ 19) from a comparable Raute community outside the study area. Internal consistency and cultural appropriateness were assessed, and revisions were made before final administration. Interviews were conducted face-to-face in the local language by the principal investigator and trained interviewers in private settings to minimize social desirability bias. Completed questionnaires were reviewed daily, and double-entry verification was performed to minimize data entry errors.

### Study variables

The primary dependent variables were: (1) current family planning use (yes/no); and (2) ever-utilization of safe abortion services (yes/no). Secondary outcomes included contraceptive method type, duration of use, perceived advantages, service source, reasons for non-use, safe abortion knowledge, service location awareness, type of procedure, and national policy awareness. Independent variables were: age, family type, residence, health service preference, age at first marriage, birth interval, number of children, number of family members, husband’s occupation, monthly household income, and literacy status. The association between family planning and safe abortion utilization was also examined bidirectionally, given the established theoretical relationship between contraceptive access and abortion demand.

### Statistical analysis

Data were entered and analyzed using IBM SPSS Statistics version 16. Descriptive statistics including frequencies, percentages, means, and standard deviations were generated for all variables. Chi-square tests examined bivariate associations between each independent variable and each dependent variable. Variables with p < 0.20 in univariate analysis were entered as candidates for multivariable logistic regression, alongside theoretically relevant covariates identified from prior literature. Variables were assessed for collinearity before inclusion in final models. Results are expressed as crude odds ratios (COR) and adjusted odds ratios (AOR) with 95% confidence intervals (CI). Exact p-values are reported for all values ≥ 0.001; values below 0.001 are reported as p < 0.001. Statistical significance was set at p < 0.05 for all tests.

## Results

### Socio-demographic and socioeconomic characteristics

Table 1 presents the full characteristics of the 192 participants. The mean age was 30.37 years (SD ± 7.34); 55.2% were aged 30 years or below. Most resided in rural areas (55.7%), and almost all (97.9%) preferred a combination of modern and traditional health services. The mean age at first marriage was 16.31 years (SD ± 2.00), with 54.2% married at or before age 16. The mean number of children was 3.17 (SD ± 1.66), and mean household size was 6.22 members (SD ± 1.98). All respondents identified as homemakers; 91.7% of husbands were laborers. Most households (67.7%) earned NPR 7,001–10,000 per month (approximately USD 47–68), and 53.1% of women were literate.

**Table 1.** Socio-demographic and socioeconomic characteristics of married Raute women of reproductive age (n = 192).

| Variable | Category | n | % |
| --- | --- | --- | --- |
| **Socio-demographic characteristics** |  |  |  |
| Age (years) | Mean ± SD: 30.37 ± 7.34 | | |
| | $\leq 30$ | 106 | 55.2 |
| | $> 30$ | 86 | 44.8 |
| Family type | Nuclear | 111 | 57.8 |
|  | Joint | 81 | 42.2 |
| Residence | Urban | 85 | 44.3 |
|  | Rural | 107 | 55.7 |
| Health service preference | Modern only | 4 | 2.1 |
|  | Modern and traditional | 188 | 97.9 |
| Age at first marriage (years) | Mean ± SD: 16.31 ± 2.00 | | |
| | $\leq 16$ | 104 | 54.2 |
| | $> 16$ | 88 | 45.8 |
| Birth interval (years) | $< 2$ | 84 | 43.8 |
|  | 2–5 | 108 | 56.3 |
| Number of children | Mean ± SD: 3.17 ± 1.66 | | |
|  | 0–2 | 84 | 43.8 |
|  | 3–5 | 87 | 45.3 |
|  | 6–10 | 21 | 10.9 |
| Number of family members | Mean ± SD: 6.22 ± 1.98 | | |
| | $\leq 6$ | 120 | 62.5 |
| | $> 6$ | 72 | 37.5 |
| **Socioeconomic characteristics** |  |  |  |
| Occupation of respondent | Homemaker | 192 | 100.0 |
| Husband's occupation | Laborer | 176 | 91.7 |
|  | Other | 16 | 8.3 |
| Monthly household income (NPR) | $\leq 7,000$ | 29 | 15.1 |
|  | 7,001–10,000 | 130 | 67.7 |
|  | 10,001–20,000 | 33 | 17.2 |
| Literacy status | Literate | 102 | 53.1 |
|  | Illiterate | 90 | 46.9 |

NPR = Nepalese Rupee; USD 1 ≈ NPR 147.68 at the time of the study; SD = standard deviation.

### Family planning and safe abortion service knowledge and utilization

Table 2 presents comprehensive data on family planning and safe abortion knowledge and utilization. More than half of respondents (53.6%) currently used family planning. Among users (n = 103), injectable contraceptives were most common (42.7%), followed by female sterilization (33.0%), implants (24.3%), and condoms (1.0%). The method mix is heavily concentrated in long-acting and permanent female-dependent methods. Concerning duration of use, 46.6% had been using their current method for less than one month, 23.3% for one month to one year, and 30.1% for more than one year. Birth spacing was the primary perceived advantage (97.1%), and 95.1% accessed services from government health facilities. Among non-users (n = 89), lack of interest was cited by 97.7%, with difficulty in access (9.0%), lack of money (2.2%), husband’s objection (1.1%), and cultural barriers (1.1%) also reported. Multiple responses were permitted.

**Table 2.** Family planning and safe abortion service knowledge and utilization among married Raute women (n = 192).

| Variable | Category | n | % |
| --- | --- | --- | --- |
| **A. Family planning service utilization** |  |  |  |
| Current family planning use | Yes | 103 | 53.6 |
|  | No | 89 | 46.4 |
| Contraceptive method used (n = 103) | Injectable (Depo-Provera) | 44 | 42.7 |
|  | Female sterilization | 34 | 33.0 |
|  | Implant | 25 | 24.3 |
|  | Condom | 1 | 1.0 |
| Duration of contraceptive use (n = 103) | <1 month | 48 | 46.6 |
|  | 1 month – 1 year | 24 | 23.3 |
|  | >1 year | 31 | 30.1 |
| Perceived advantage of family planning (n = 103) | Birth spacing | 100 | 97.1 |
|  | STI protection | 2 | 1.9 |
|  | Better maternal and child health | 1 | 1.0 |
| Source of services (n = 103) | Government health facility | 98 | 95.1 |
|  | Non-government/private facility | 5 | 4.9 |
| Reasons for non-use (n = 89) | Lack of interest | 87 | 97.7 |
|  | Difficulty in access | 8 | 9.0 |
|  | Lack of money | 2 | 2.2 |
|  | Husband's objection | 1 | 1.1 |
|  | Cultural barrier | 1 | 1.1 |
| **B. Safe abortion service utilization** |  |  |  |
| Knowledge of safe abortion services | Yes | 118 | 61.5 |
|  | No | 74 | 38.5 |
| Knowledge of service location (n = 118) | Government health facility | 114 | 96.6 |
|  | Non-government/private facility | 4 | 3.4 |
| Utilization of safe abortion services | Yes | 16 | 8.3 |
|  | No | 176 | 91.7 |
| Type of procedure received (n = 16) | Medical abortion | 10 | 62.5 |
|  | Surgical abortion | 6 | 37.5 |
| Knowledge of Nepal's national safe abortion policy | Yes | 21 | 10.4 |
|  | No | 171 | 89.6 |
| Knowledge of specific policy provisions (n = 21) | Available on request up to 12 weeks | 14 | 66.7 |
|  | Up to 18 weeks in rape/incest cases | 4 | 19.0 |
|  | Anytime when mother's life is at risk | 3 | 14.3 |

Percentages for method type, duration, perceived advantages, and service source are from current users (n = 103). Reasons for non-use are from non-users (n = 89); multiple responses permitted. Service location percentages are from women with safe abortion knowledge (n = 118). Procedure type percentages are from women who utilized safe abortion services (n = 16). Policy provision percentages are from women with policy awareness (n = 21). STI = sexually transmitted infection.

Regarding safe abortion, 61.5% of women reported knowledge of services, of whom 96.6% correctly identified government facilities as the source of care. Despite this, only 8.3% had ever accessed safe abortion services. Among those who had utilized services (n = 16), medical abortion was more common (62.5%) than surgical abortion (37.5%). Knowledge of Nepal’s national safe abortion policy was critically low at 10.4%. Among the 21 women with policy awareness, 66.7% knew abortion is available on request up to 12 weeks, 19.0% were aware of provisions for rape or incest up to 18 weeks, and 14.3% knew it is available at any gestation when the mother’s life is at risk.

### Factors associated with family planning utilization

Table 3 presents chi-square and logistic regression results for current family planning utilization. No socio-demographic or socioeconomic variable was significantly associated with family planning use in bivariate or adjusted analysis. A borderline association was observed for birth interval (p = 0.082), with women with intervals of two to five years more likely to use family planning than those with shorter intervals. The association between safe abortion utilization and family planning use reached statistical significance in bivariate analysis (p = 0.008). In the multivariable logistic regression model, the sole significant independent predictor of family planning use was non-utilization of safe abortion services (AOR = 4.275; 95% CI: 1.145–15.954; p = 0.030), indicating that women who had not accessed abortion services were over four times more likely to be current family planning users after adjustment for age, residence, and literacy.

**Table 3.**
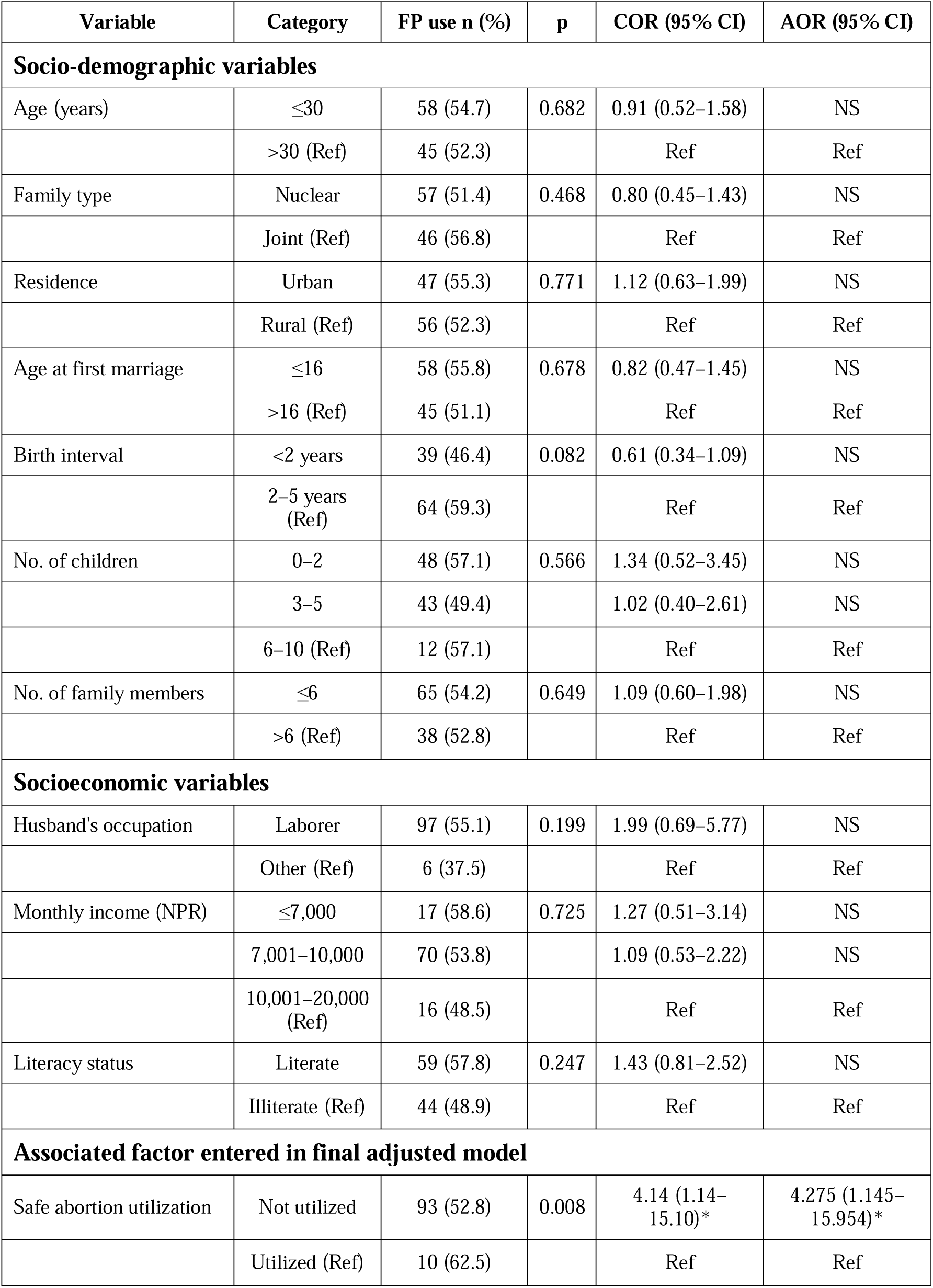
Factors associated with current family planning utilization: chi-square and logistic regression analysis (n = 192).

### Factors associated with safe abortion service utilization

Table 4 presents chi-square and logistic regression results for safe abortion utilization. Two socio-demographic variables reached significance in bivariate analysis. Age was significantly associated (p = 0.003): 13.2% of women aged 30 years or below utilized services versus 2.3% of older women (COR = 6.391; 95% CI: 1.411–28.956). Residence was also significant (p = 0.039): urban women showed a 12.9% utilization rate versus 4.7% for rural women (COR = 3.032; 95% CI: 1.011–9.098). In the adjusted model, both age (AOR = 4.552; 95% CI: 0.890–23.273; p = 0.071) and urban residence (AOR = 2.933; 95% CI: 0.916–9.394; p = 0.068) were attenuated to non-significance, consistent with limited statistical power from the small number of outcome events (n = 16). No socioeconomic variable reached significance in either analysis, though a trend toward higher utilization with increasing household income was observed.

**Table 4.** Factors associated with safe abortion service utilization: chi-square and logistic regression analysis (n = 192).

| Variable | Category | SA use n (%) | p | COR (95% CI) | AOR (95% CI) |
| --- | --- | --- | --- | --- | --- |
| **Socio-demographic variables** |  |  |  |  |  |
| Age (years) | ≤30 | 14 (13.2) | 0.003 | 6.39 (1.41–28.96)* | 4.55 (0.89–23.27) |
|  | >30 (Ref) | 2 (2.3) |  | Ref | Ref |
| Family type | Joint | 10 (12.3) | 0.086 | 2.47 (0.87–7.02) | NS |
|  | Nuclear (Ref) | 6 (5.4) |  | Ref | Ref |
| Residence | Urban | 11 (12.9) | 0.039 | 3.03 (1.01–9.10)* | 2.93 (0.92–9.39) |
|  | Rural (Ref) | 5 (4.7) |  | Ref | Ref |
| Age at first marriage | ≤16 | 10 (9.6) | 0.450 | 1.44 (0.51–4.10) | NS |
|  | >16 (Ref) | 6 (6.8) |  | Ref | Ref |
| Birth interval | <2 years | 5 (6.0) | 0.292 | 0.55 (0.18–1.64) | NS |
|  | 2–5 years (Ref) | 11 (10.2) |  | Ref | Ref |
| No. of children | 0–2 | 8 (9.5) | 0.341 | 0.99 (0.33–2.93) | NS |
|  | 3–5 | 8 (9.2) |  | 1.01 (0.35–2.98) | NS |
|  | 6–10 (Ref) | 0 (0.0) |  | Ref | Ref |
| No. of family members | ≤6 | 12 (10.0) | 0.196 | 1.88 (0.59–5.98) | NS |
|  | >6 (Ref) | 4 (5.6) |  | Ref | Ref |
| **Socioeconomic variables** |  |  |  |  |  |
| Husband's occupation | Laborer | 14 (8.0) | 0.529 | 0.57 (0.12–2.66) | NS |
|  | Other (Ref) | 2 (12.5) |  | Ref | Ref |
| Monthly income (NPR) | ≤7,000 | 1 (3.4) | 0.225 | 0.42 (0.05–3.35) | NS |
|  | 7,001–10,000 | 10 (7.7) |  | 0.99 (0.28–3.46) | NS |
|  | 10,001–20,000 (Ref) | 5 (15.2) |  | Ref | Ref |
| Literacy status | Literate | 11 (10.8) | 0.191 | 2.06 (0.70–6.04) | NS |
|  | Illiterate (Ref) | 5 (5.6) |  | Ref | Ref |

COR = crude odds ratio; AOR = adjusted odds ratio; CI = confidence interval; Ref = reference category; NS = not statistically significant ( $p \geq 0.20$ in univariate analysis or $p \geq 0.05$ in adjusted model); SA = safe abortion. \* $p < 0.05$ in bivariate analysis. The final adjusted model included age and residence. Attenuation reflects limited statistical power ( $n = 16$ events). $p$ -values from two-sided chi-square tests. NPR = Nepalese Rupee.

## Discussion

This census-based study provides the first population-level quantitative evidence on family planning and safe abortion knowledge and utilization among married Raute women of reproductive age. The findings reveal a community in transition: meaningful progress in contraceptive coverage has been achieved, yet profound gaps persist in method diversity, safe abortion policy literacy, and reproductive autonomy.

### Family planning utilization

The family planning prevalence of 53.6% exceeds Sudurpaschim Province’s reported rate of 47% and approaches the national figure of 57% [12], a notable achievement given the community’s nomadic lifestyle and historical resistance to external interventions. This suggests that sustained outreach by government health workers, female community health volunteers (FCHVs), and NGO programs has achieved meaningful penetration. However, nearly half of women remain non-users, and the method mix is severely imbalanced. Injectable contraceptives, sterilization, and implants account for virtually all use, while condom use is effectively absent at 1.0%. This mirrors findings from the Mru indigenous community of Bangladesh [23] and tribal women of West Bengal, India [24], where male-controlled methods are consistently underrepresented across South and Southeast Asian indigenous communities. The near-absence of condoms means family planning remains a female-only responsibility in the Raute community, with male partners structurally disengaged, a pattern documented broadly across Nepal’s marginalized groups [13, 14]. The dominance of “lack of interest” as the reason for non-use (97.7%) likely masks deeper barriers. Prior Nepali research consistently documents that stated disinterest conceals fear of side effects, partner opposition, internalized cultural norms, and the absence of counseling to address these concerns [25, 26]. Only 1.1% explicitly cited cultural barriers, not because culture is absent as a constraint but because cultural norms are so thoroughly internalized that they present as personal preference.

### Safe abortion knowledge and the knowledge-to-practice gap

The knowledge-to-practice gap in safe abortion is the most critical finding of this study. While 61.5% of women knew about safe abortion services and 96.6% correctly identified government facilities as the source of care, only 8.3% had ever accessed them — a gap exceeding 50 percentage points. National policy awareness at 10.4% is alarmingly low, comparing unfavorably to 38.7% reported among general-population women in prior national studies [16]. Nepal’s abortion law has been in effect since 2002; that the majority of Raute women remain unaware of their legal right to abortion more than two decades later is a systematic failure of health communication and outreach. The NDHS trend analysis shows national policy awareness increased by 6.4 percentage points between survey cycles [12], yet the Raute community has not benefited from this progress. The significant bivariate associations between younger age and urban residence with safe abortion use align with prior Nepali evidence [27]: younger women face greater fertility pressure and are more likely to have encountered health information, while urban women benefit from proximity to facilities. Both associations were attenuated after adjustment, reflecting limited statistical power rather than a genuine absence of effect. Beyond structural access, abortion-seeking decisions are constrained by stigma, fatalism, and community surveillance that characterize health behavior in a small, closely-knit nomadic group [21, 28].

### The inverse association between family planning and safe abortion use

The finding that non-utilization of safe abortion services independently predicted current family planning use (AOR = 4.275; 95% CI: 1.145–15.954; p = 0.030) is analytically important. Women who had not accessed abortion services were over four times more likely to use family planning, suggesting that contraceptive use and abortion use represent substitutable strategies for managing unintended pregnancies: women who use contraception prevent unintended pregnancies and therefore have less need for abortion, while non-users may rely on abortion as a fallback. This relationship is well established across low- and middle-income settings [5, 15, 28] and the present data provide the first community-level evidence of this pattern among a nomadic indigenous population. The implication is direct: strengthening family planning access is the most efficient strategy to simultaneously reduce unintended pregnancies and diminish reliance on abortion services in this community.

### Strengths and limitations

The primary strength of this study is the census design, which enrolled all eligible married Raute women, eliminated selection bias, and produced the first population-level reproductive health data for this community. The simultaneous assessment of family planning and safe abortion enables examination of their relationship at a community level. Limitations include the cross-sectional design, which precludes causal inference. Social desirability bias may lead to underreporting of abortion utilization; the true rate may be higher than 8.3%. The small number of safe abortion events (n = 16) substantially limits regression model power for that outcome, producing wide confidence intervals. The study was conducted in a single municipality and may not fully generalize to Raute populations in other districts. Qualitative inquiry is needed to explore the cultural dynamics and social constraints that underlie the observed patterns.

### Programmatic and policy implications

Parshuram Municipality and Sudurpaschim Province health authorities should establish mobile reproductive health outreach tailored to Raute seasonal movement patterns, integrating family planning supply, safe abortion referral, and legal literacy communication in a single service package. Male partner engagement is required to shift contraceptive decision-making from a female-only domain to a shared responsibility. Safe abortion legal literacy must be communicated through oral and visual channels in the local language, given 46.9% illiteracy. FCHVs require training and logistical support adapted to nomadic contexts. At the national level, nomadic indigenous communities must be explicitly recognized as priority populations in Nepal’s Family Planning Strategy, Safe Abortion Policy, and National Health Policy, with dedicated service access monitoring.

### Conclusions

This study provides the first population-level evidence on family planning and safe abortion knowledge and utilization among married Raute women of reproductive age. Family planning uptake at 53.6% reflects real progress, yet method diversity is severely limited, male-controlled contraceptives are essentially absent, and safe abortion services are accessed by only 8.3% of women despite 61.5% having knowledge of their existence. Safe abortion policy awareness at 10.4% — nearly 30 percentage points below the general population — represents a critical legal literacy deficit more than two decades after Nepal legalized abortion. The inverse association between contraceptive use and abortion service utilization provides empirical justification for prioritizing family planning access as the primary strategy to reduce unintended pregnancy. Mobile reproductive health outreach, male partner engagement, legal literacy programs in the local language, and explicit recognition of nomadic indigenous communities in national reproductive health policy are all essential and actionable priorities.

## Acknowledgements

The authors sincerely thank all married women of the Raute community of Parshuram Municipality who participated in this study. Gratitude is extended to the health section and municipal executive office of Parshuram Municipality for granting permission and facilitating community access. The Department of Public Health, Manmohan Memorial Institute of Health Sciences is acknowledged for institutional support. The contributions of local health post staff and female community health volunteers who facilitated data collection are gratefully acknowledged.

## Author contributions

**Conceptualization:** MJ, SK.

**Data curation:** MJ, AB.

**Formal analysis:** MJ, SK,

**Investigation:** MJ, AB.

**Methodology:** MJ, SK.

**Project administration:** MJ.

**Supervision:** SK.

**Validation:** SK.

**Writing – original draft:** MJ, AS.

**Writing – review and editing:** SK, AB, AS, MT

## Data availability

The datasets generated during this study are not deposited in a public repository because: (1) data were collected under ethical approval conditions requiring participant confidentiality; (2) the census design in a small, identifiable community of 419 individuals means that even de-identified records carry a meaningful risk of individual re-identification; and (3) this restriction was explicitly agreed with participants during informed consent. Summary-level data supporting all findings reported in this manuscript are presented in Tables 1–4 within the article. Individual-level de-identified data may be made available from the corresponding author on reasonable written request, subject to review and approval by the Institutional Review Committee of Manmohan Memorial Institute of Health Sciences.

## Supporting information

**S1 Questionnaire.** English translation of the structured data collection instrument used in this study. The original instrument was administered in the local language.

**S2 Ethics approval.** Institutional Review Committee approval letter, Manmohan Memorial Institute of Health Sciences (MMIHS-IRC 886, 1 September 2022).

**S3 STROBE checklist.** Completed STROBE checklist for cross-sectional studies.

